# Prevalence and etiology of strabismus in Down syndrome: A systematic review and meta-analysis with a focus on ethnic differences in the esotropia/exotropia ratio

**DOI:** 10.1101/2024.11.28.24318156

**Authors:** Christopher S. von Bartheld, Avishay Chand, Lingchen Wang

## Abstract

**Purpose:** We sought to determine the prevalence of strabismus and the esotropia/exotropia ratio in Down syndrome. Wide ranges of an increased strabismus prevalence have been reported and it is unclear by how much esotropia exceeds exotropia in people with Down syndrome.

**Methods:** We compiled in a systematic review and meta-analysis results of over 100 studies that report the strabismus prevalence and ratio of esotropia/exotropia in cohorts of Down syndrome. We calculated the pooled global prevalence and established the geographical distribution of the strabismus prevalence and the esotropia/exotropia ratio.

**Results:** The ethnically-adjusted global prevalence of strabismus in Down syndrome is 30.2%. In subjects 15 years and older, the global prevalence is 53.2%, and the lifetime prevalence is 51.0%. In populations which normally have more esotropia than exotropia (e.g., Caucasians), Down syndrome subjects have a further increased bias towards esotropia. In populations which normally have more exotropia (e.g., West Africans, Asians and Hispanics), Down syndrome subjects have a significantly lower esotropia/exotropia ratio (3.21) than reported in Caucasians with Down syndrome (9.98).

**Conclusion:** Worldwide, about 1.81 million people with Down syndrome have strabismus: 1.42 million of them have esotropia, and 0.37 million have exotropia. Differences in the esotropia/exotropia ratio between ethnicities point to the orbital anatomy as a major contributing factor to the etiology of strabismus in Down syndrome. The narrow-set eyes (reduced orbital width) in Down syndrome favor esotropia over exotropia, especially in Caucasians, thus explaining why Down syndrome patients from different ethnicities have different prevalences of esotropia and exotropia.

## Introduction

Since the discovery of an increased frequency of strabismus in people with Down syndrome,^1–4^ many authors have reported the prevalence of strabismus. Review articles and primary research articles have reported diverse ranges of the strabismus prevalence, from 1.9% to 100% (Supplemental Table 1). Arbitrary selections of studies led to the variety of the reported ranges in review articles, and the true prevalence of strabismus in Down syndrome has remained elusive. In addition, reports of the ratio of esotropia to exotropia in Down syndrome have been widely divergent. Nearly half of the European studies in the 20^th^ century reported exclusively esotropia, and no exotropia in their cohorts, while most of the studies from Asia and Africa reported at least 25% of the strabismus cases to be exotropia. Some of the differences may be due to Eurocentric bias and neglect of studies published in non-English languages. Our review includes studies published in thirteen languages other than English and is the first review that systematically compiles relevant studies. The previous most thorough reviews considered only a small fraction (13 to 31) ^5–9^ of the 142 available reports on the prevalence of strabismus in Down syndrome.

Several authors reported that the strabismus in Down syndrome children develops at a significantly later age than in the normal population,^4–5,10–20^ indicating that the lifetime prevalence of strabismus in Down syndrome may be higher than the global prevalence. Probably because of the late onset of the acquired strabismus in Down syndrome, a remarkably low percentage of amblyopia was noted in many studies.^5,15,21–31^ Some authors reported ethnic differences in the frequency of esotropia vs exotropia in Down syndrome,^14,17,32–39^ but possible underlying mechanisms were not explored.

The etiology of strabismus in Down syndrome has remained enigmatic and controversial. Several potential causes of strabismus in Down syndrome were discussed, including the narrow orbital width in Down syndrome, opacity of the lens, muscle hypotonia, refractive errors with lack of normally occurring emmetropization, accommodation weakness, retinal abnormalities, visual cortex abnormalities, and various combinations of the above conditions.^5,17–19,21,37,40–51^ The etiology of strabismus in Down syndrome differs substantially from that in the normal population.^49–51^ Yet, a comprehensive review of the etiology of strabismus in Down syndrome is lacking.

In our systematic review and meta-analysis,^52^ we provide information about the true global prevalence of strabismus in people with Down syndrome. We estimate global numbers of esotropia and exotropia cases that take into account the Eurocentric bias due to the large majority of studies examining Caucasians of European ancestry, necessitating the calculation of an ethnically-adjusted global prevalence. We provide a geographic world map of the esotropia/exotropia ratio in Down syndrome, and we review and synthesize opinions and arguments about the etiology of strabismus in Down syndrome.

## Materials and Methods

### Search Strategy

For our systematic review of the literature, we adhered to the PRISMA guidelines.^53^ Reports of studies were identified through a search of Google Scholar and PubMed, with unrestricted years. We used the keywords “Down syndrome”, “trisomy 21”, “strabismus”, “esotropia”, and “exotropia” in Google Scholar, and “Down syndrome” or “trisomy 21” and “strabismus”, as well as “Down syndrome” or “trisomy” and “squint” in PubMed. Only English terms were used for the search strategy, but we retrieved 29 studies that were published in languages other than English, because they had an English title and/or abstract, or were cited in relevant studies. Studies published in languages besides English included German (5 studies), Turkish (5), French (3), Portuguese (3), Spanish (3), Norwegian (2), Polish (2), Chinese (1), Czech (1), Italian (1), Japanese (1), Russian (1), and Swedish (1). References cited in eligible articles were examined to identify additional relevant studies. Titles were screened, and when potentially relevant, the abstract was evaluated to decide whether full-text should be obtained to verify eligibility (Fig. 1). We failed to obtain an abstract or full-text in 3 of 817 sources.

**Fig. 1.**
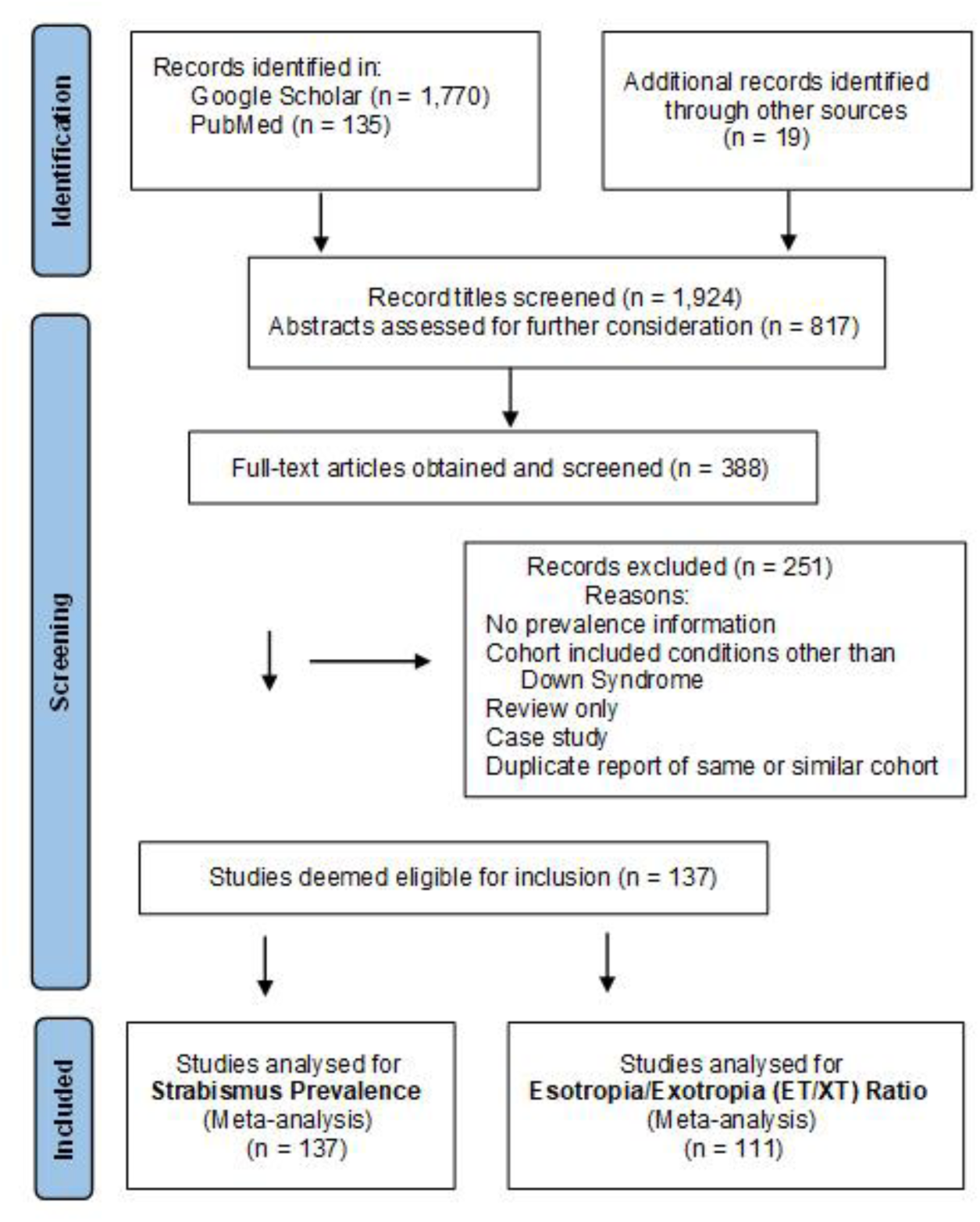
Flowchart of the Literature Search.

### Inclusion/Exclusion Criteria

To be eligible for inclusion in our systematic review, studies had to report the numerical prevalence of strabismus in humans with Down syndrome (non-human primates were not considered)^54^ and/or provide the ratio of esotropia vs exotropia in a Down syndrome cohort. We excluded reviews only, case reports, and abstracts at meetings when later published as a peer-reviewed paper. Sorted by geography, we included 55 studies on Caucasians in Europe,^1–5,11,28,33–34,40,44,47–49,55–94^ 29 studies from North America or Australia,^21,23,29,31,42,95–118^ 18 from the Middle East,^8–9,12–13,26–27,45,119–129^ 8 from Latin America,^10,25,30,130–134^ 13 from East Asia,^14,32,35,135–144^ 12 from South Asia,^17–18,20,145–153^ and 7 from Africa or on Africans,^154–160^ for a total of 142 eligible studies (Supplemental Table 1). For the final analyses, we excluded 5 studies that reported on duplicate cohorts.

### Data Extraction and Analyses

Data were extracted by using pre-designed tables, including year of publication, first author name, country, geographic region, age range, cohort size, number of cases of strabismus, and the type of strabismus: horizontal vs. vertical, and among the horizontal strabismus, how many cases of esotropia, how many cases of exotropia and the esotropia/exotropia ratio. Cases of microtropia and paralytic strabismus were not included. When gender distribution in the cohort was reported, we compiled such information, and also information on gender in strabismus cases. The percentage of strabismus cases was calculated from the number of cases examined per cohort. We conducted subgroup analyses between continents and ethnicities. Because of ethnic differences between populations in the ratio of esotropia/exotropia, the prevalence for Caucasians and non-Caucasians was estimated separately and weighted by population size to generate a global estimate of the esotropia/exotropia ratio and numbers of esotropia and exotropia cases. This was also necessary to prevent bias: a large majority of available studies examined people of European ancestry. We had sufficient data for Caucasians and populations from the Middle East to assess generational (longitudinal) trends. We also performed subgroup analyses for different age ranges and estimated the lifetime prevalence of strabismus in Down syndrome.

### Statistical Analyses

A major purpose of our meta-analysis was to generate a more precise and reliable estimate of the prevalence of strabismus among people with Down syndrome and to infer the global number of Down syndrome cases with strabismus. A second purpose was to analyze the esotropia/exotropia (ET/XT) ratio to determine potential differences between ethnicities. For this analysis, three of the studies were grouped by ethnicity rather than geography because the ethnicity of the subjects differed from the geography (Supplementary Table 1).^10,116,159^ Pooled analyses were performed for strabismus prevalence in Down syndrome and the ET/XT ratio. The prevalence was subjected to a Freeman–Tukey double arcsine transformation, and the ET/XT ratio was log-transformed. The heterogeneity among studies was evaluated by Cochran’s Q test, the I^2^ index, and predictive intervals.^161–162^ The random-effect models were used to conservatively diminish the heterogeneity between studies.^162^ The study weights were obtained based on the DerSimonian-Laird method.^162^ Subgroup pooled analyses were conducted by region/ethnicity, separately for prevalence and for the ET/XT ratio, to assess differences between Caucasians and non-Caucasians. When calculating the ET/XT ratio, a continuity correction of 0.5 was applied to studies with zero XT cases by adding 0.5 to both ET and XT cases to avoid division by zero.^163^ Meta-regression analyses were performed to test associations between independent variables (age, gender, ethnicity, year of publication) and response variables (prevalence, ET/XT ratio). The risk of publication bias was evaluated using funnel plots and Egger’s test (Supplemental Figs. 1, 3).^164^ The significance level was set to 0.05. All meta-analyses were performed using the Stata SE 16.0 software (StataCorp, TX, USA).

## Results

We included in our analyses 137 of the total 142 eligible studies (after removal of 5 duplicate publications of largely the same cohort). Their geographical distribution and cohort sizes are depicted in Figure 2A. Cohort sizes varied from 3 to 1,539, with a mean cohort size of 122.5 and a total number of 16,781 subjects in the cohorts. There was no publication bias for Caucasians, but there was bias for Non-Caucasians based on funnel plots (Supplemental Fig. 1A,B). 60.6% of all eligible studies (83/137) and 65.3% of all subjects in the cohorts were on Caucasians or mostly Caucasians, while Asians and Africans were underrepresented: only 16.1% of subjects, combined, were Asians and Africans. The world map indicates that Caucasians have a larger strabismus prevalence than other ethnicities (Fig. 2A). We therefore tested in a subgroup analysis whether there was a significant difference in prevalence between Caucasians and other ethnicities or geographical areas. Caucasians have a 39.0% prevalence, while Non-Caucasians have a 28.7% prevalence – a significant difference (p=0.001). The Forest plot for the prevalence of strabismus in Down syndrome in Caucasians and Non-Caucasians is shown in Fig. 3A,B. When adjusted for population size of ethnicities (1.2 billion Caucasians, 0.5 billion Middle Easterners, 1.9 billion South Asians, 2.4 billion East Asians, 1.3 billion Africans and 0.7 billion Hispanics), the global prevalence of strabismus in Down syndrome was 30.2% (95% confidence interval, CI, = 29.2-31.3%). The ethnically-adjusted pooled prevalence of strabismus in Down syndrome allows us to estimate the total number of people with Down syndrome – worldwide – who have strabismus. Based on estimates of 6 million people worldwide with Down syndrome,^165^ we conclude that 1.81 million have strabismus, with 1.42 million having esotropia and 0.37 million having exotropia (see below).

**Fig. 2.**
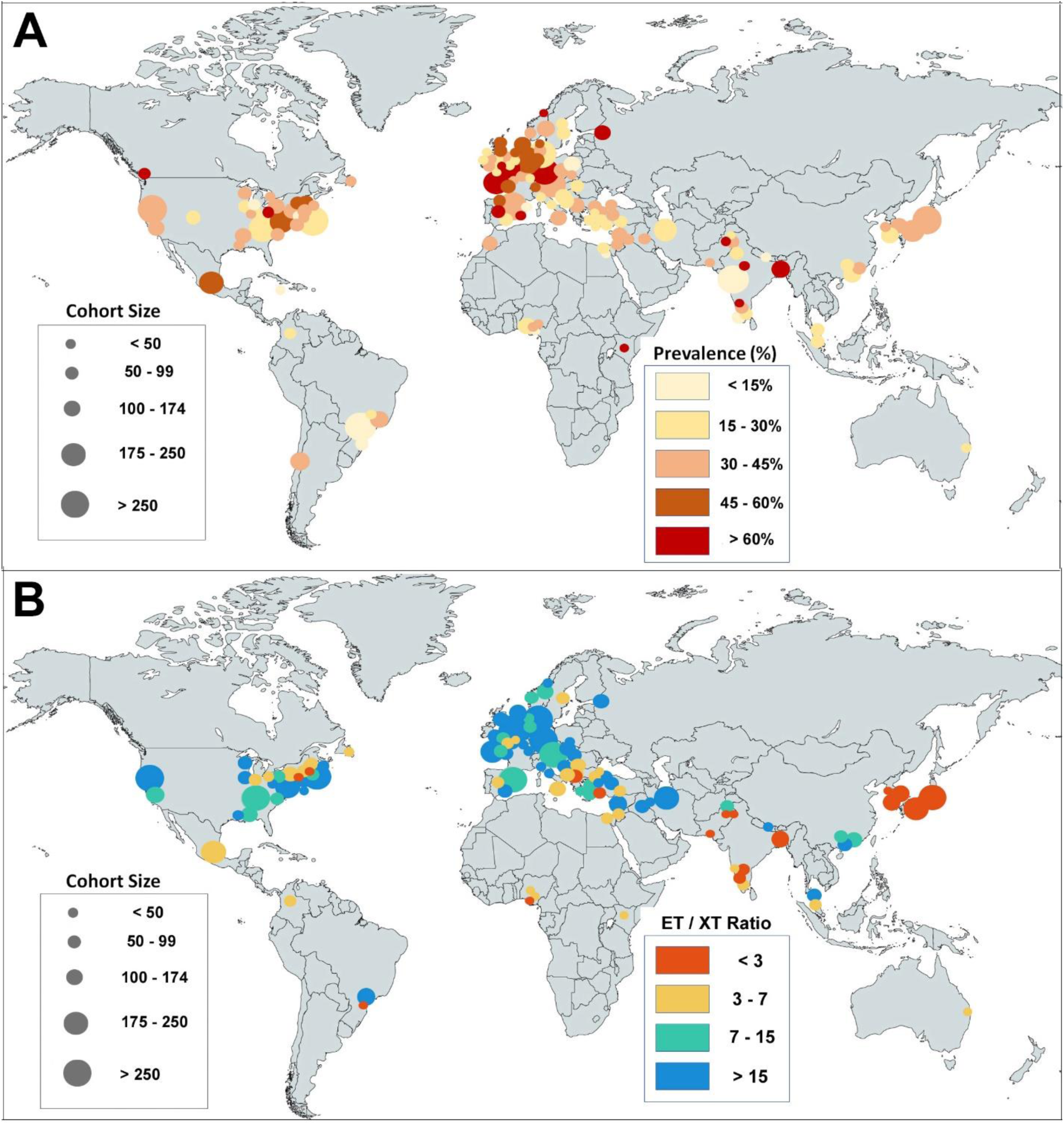
**A,B.** Distribution of studies reporting the prevalence of strabismus in Down syndrome (**A**) and distribution of studies reporting the esotropia / exotropia (ET/XT) ratio in Down Syndrome (**B**). The prevalence and ET/XT ratio are indicated in a heat map. Cohort Sizes are indicated by the size of the circles.

**Fig. 3.**
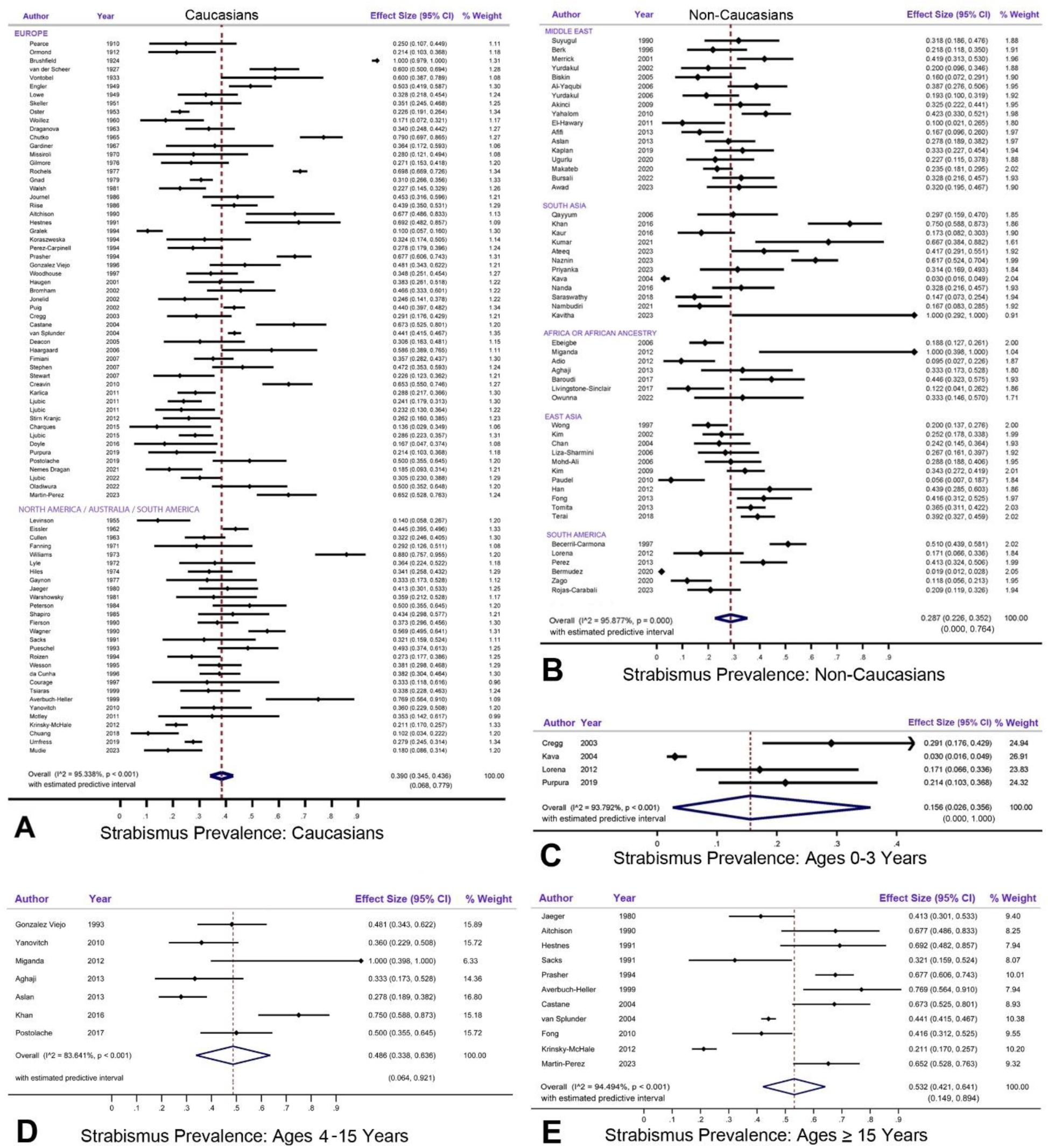
**A-D.** Forest Plots show the strabismus prevalence in Down syndrome. CI, confidence interval; I^^2^ indicates the level of heterogeneity. **A**. In Caucasians. **B**. In Non-Caucasians (Middle East, South Asia, Africa, East Asia, South America). **C**. Ages 0-3 years. **D**. Ages 4-15 years. **E**. Ages 15 years and older.

AGE. Studies examined strabismus prevalence for different age groups of Down syndrome. The prevalence was low in the first three years and increased at about 4 years of age.^4,9–17,19–20^ The prevalence of strabismus in cohorts of subjects below 3 years of age was 15.6% (CI = 2.6-35.6%, Fig. 3C), it was 48.6% (CI = 33.8-63.6%) for ages 4-15 years (Fig. 3D), and 53.2% (CI = 42.1-64.1%) for subjects older than 15 years (Fig. 3E). Adults (18 years and older, n=8 studies; n=2,045 subjects) had a 51.0% (CI = 38.0-63.8%) prevalence of strabismus – considered the lifetime prevalence. We estimate that the current global number of Down syndrome people with strabismus is 1.81 million, but that 3.19 million will develop strabismus during their lifetime.

GENDER. Among the 137 studies, 78 reported the gender distribution in the cohort (total number of subjects=11,036): the average sex ratio in the time period from 1910 to 2024 was 1.21 males per female, which is nearly identical to a previous review (1.22 in the 1990s).^166^ Only 7 studies (cohort size=793)^1,4,9,27,71–72,96^ reported the gender distribution also among strabismus cases. The pooled prevalence of strabismus in males was 36.2% (CI = 22.3-51.2%) and in females 38.6% (CI = 25.7-52.2%), which is no significant difference (p=0.866, Fig. 4B).

**Fig. 4.**
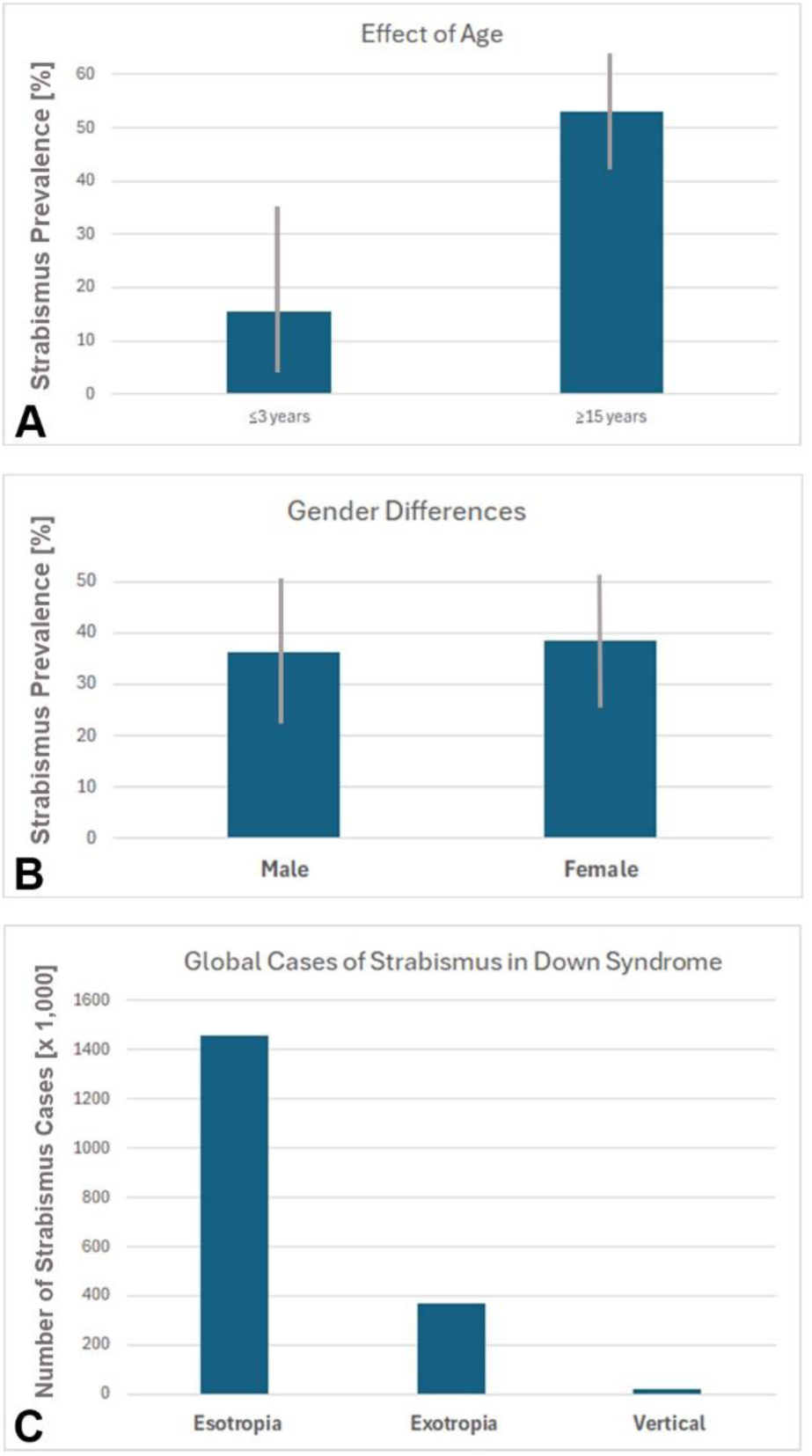
**A-C.** Graphs showing significant differences in strabismus prevalence by age **(A)** <3 years and 15 years and older, no difference in prevalence by gender **(B),** and the estimated number of people with Down syndrome having esotropia, exotropia and vertical deviations **(C).** Error bars in A,B = 95% confidence intervals.

TYPES OF DEVIATION. Among the 137 studies that reported the prevalence of strabismus in Down syndrome, 111 distinguished the prevalence of esotropia and exotropia, and among these 111 studies, 31 reported the number of the (relatively rare) cases of vertical deviations. Based on these 31 studies (3,991 subjects, Supplementary Table 1), we calculated a prevalence of 1.0% for vertical strabismus in Down syndrome (95% CI = 0.4-1.8%). Globally, this amounts to 18,444 Down syndrome cases with vertical deviation (Fig. 4C).

ESOTROPIA/EXOTROPIA RATIO. We compiled the geographic distribution of the prevalence of esotropia vs. exotropia, as indicated in 111 studies that reported these data (Fig. 2B). It is apparent in the world map that the esotropia/exotropia ratio is higher in Europe and North America than in the Middle East, Africa, Latin America, and most of Asia. We therefore analyzed the esotropia/exotropia ratio separately for Caucasians vs. non-Caucasians (Fig. 5A-C). The esotropia/exotropia ratio in Caucasians (including Middle Easterners) was 9.981 (95% CI = 7.960-12.514), while in Non-Caucasians it was 3.206 (95% CI = 2.421-4.246). We estimated the number of people with Down syndrome who have esotropia or exotropia, assuming for these estimates a similar Down syndrome prevalence in different ethnicities.^167–170^ The ethnicity-adjusted estimated numbers for Down syndrome people with esotropia are 346,000 of European ancestry (including Middle Easterners), and 1.077 million non-Caucasians, for a total of 1.423 million esotropes. The numbers for Down syndrome people with exotropia are 34,700 for European ancestry (including Middle Easterners), and 336,000 for non-Caucasians, for a total of 370,700 exotropes.

**Fig. 5.**
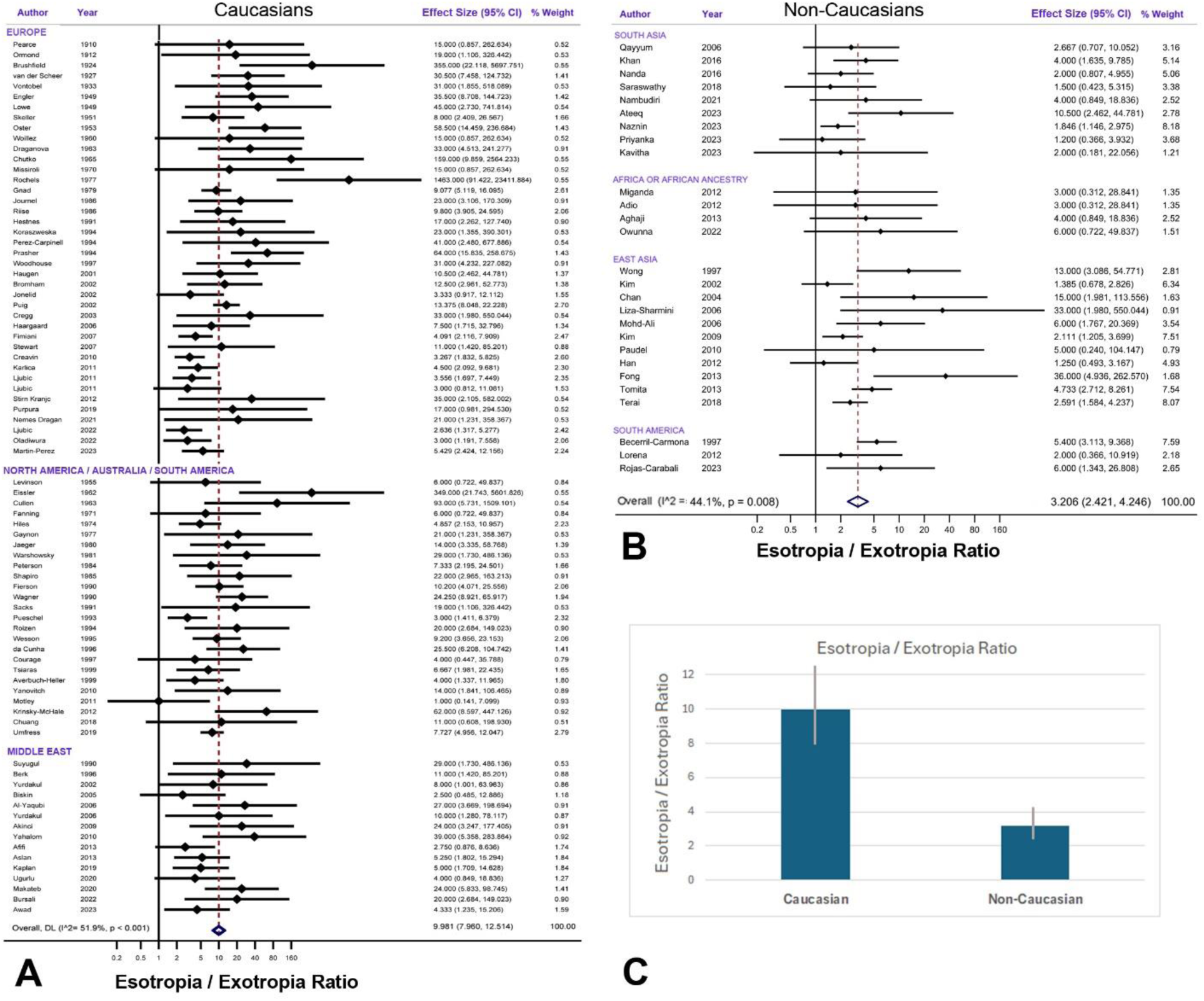
**A-C.** Forest plots of the Esotropia/Exotropia Ratio in Down syndrome for Caucasians and populations from the Middle East **(A),** and Asians, Africans and Hispanics **(B).** CI, confidence interval; I^^2^ indicates the level of heterogeneity. Bar graph shows the esotropia/exotropia ratio in Caucasians vs Non-Caucasians with Down syndrome **(C),** with error bars indicating the 95% confidence intervals.

TRENDS. There were no significant longitudinal trends in strabismus prevalence in Caucasians or in Middle Easterners (p=0.371, and 0.699, respectively, Supplemental Figure 2A-C). The strabismus prevalence appears to be stable over generations.

However, the ET/XT ratio showed a significant decrease over time in Europe (p<0.001), but there was no such trend in North America or in the Middle East (p=0.339, p=0.442, respectively, Supplemental Fig. 4A-C), possibly due to a lack of data from the first half of the 20^th^ century.

## Discussion

Our systematic review and meta-analysis provide a resolution to the conflicting reports and diverse ranges of strabismus prevalence in Down syndrome reported previously (Table 1).^5–9,13–17,19,21–24,27–28,32–33,36,43,51,59,64,67,72,76,88,99–101,104,107,121,123,125,131–132,134–136,140,156,171–174^ We can now estimate the global prevalence of such strabismus, take into account Eurocentric biases, define ethnic differences in the esotropia/exotropia ratio, resolve the age-dependence of strabismus prevalence, and answer questions about possible gender differences. It is currently controversial whether strabismus in Down syndrome associates with an increased degree of intellectual disability.^21,29,45,82,138,175–176^ We confirm that the onset of strabismus in Down syndrome typically occurs later than in the normal population. The second half of our Discussion reviews previous and current thinking about the still mysterious etiology of strabismus in Down syndrome, and concludes that ethnic differences in the esotropia/exotropia ratio help to better understand what causes strabismus in Down syndrome.

**Table 1.**
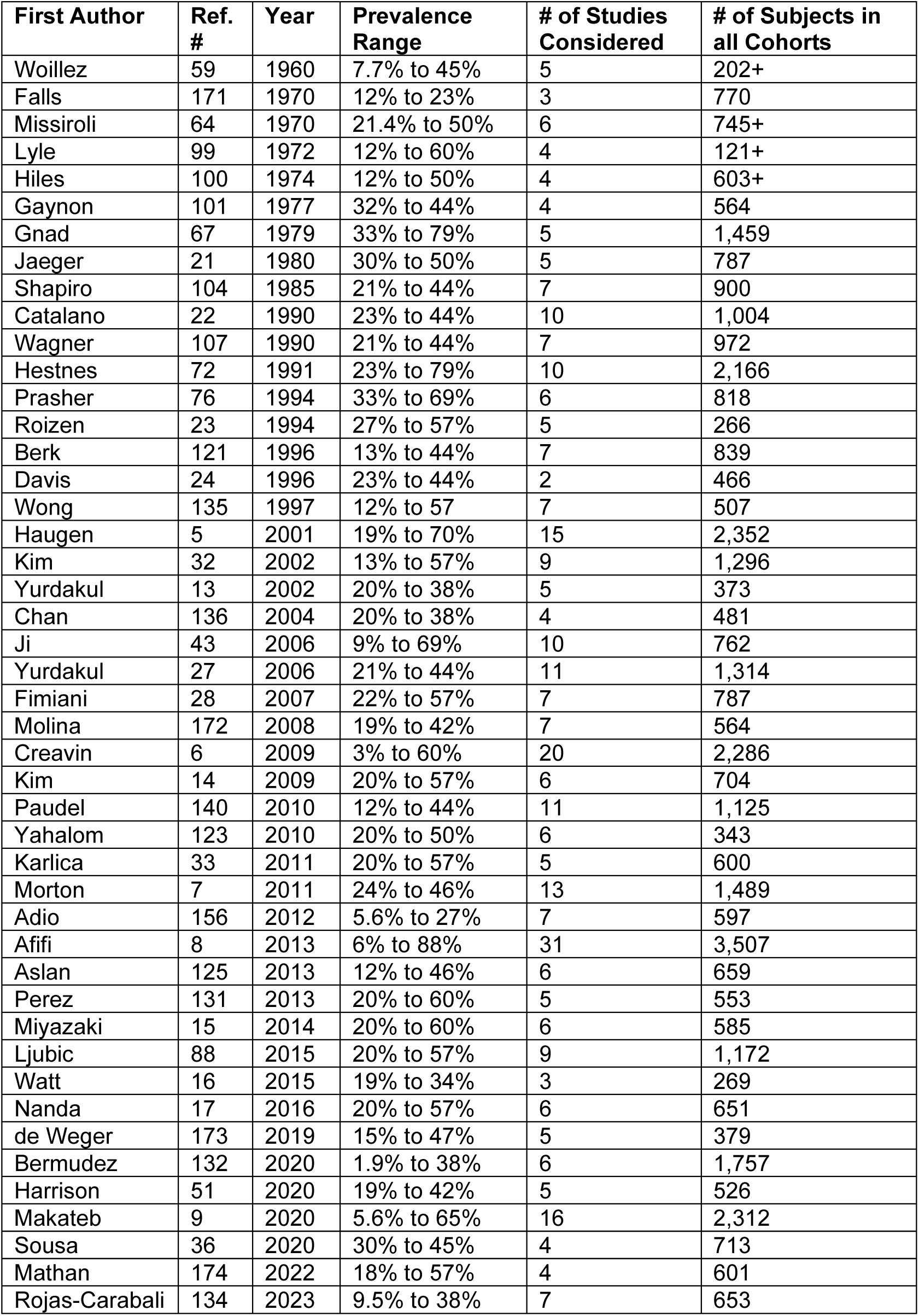
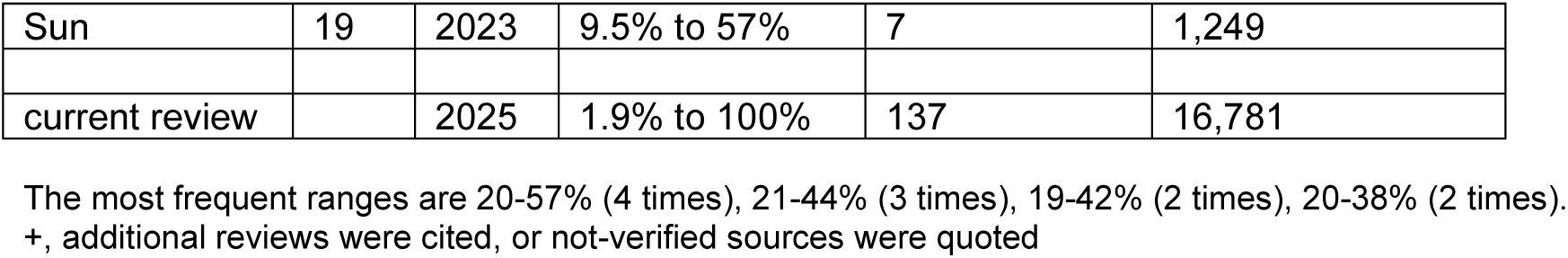
Forty different ranges of strabismus prevalence in Down syndrome according to previous authors.

| First Author | Ref. # | Year | Prevalence Range | # of Studies Considered | # of Subjects in all Cohorts |
| --- | --- | --- | --- | --- | --- |
| Wuillez | 59 | 1960 | 7.7% to 45% | 5 | 202+ |
| Falls | 171 | 1970 | 12% to 23% | 3 | 770 |
| Missiroli | 64 | 1970 | 21.4% to 50% | 6 | 745+ |
| Lyle | 99 | 1972 | 12% to 60% | 4 | 121+ |
| Hiles | 100 | 1974 | 12% to 50% | 4 | 603+ |
| Gaynon | 101 | 1977 | 32% to 44% | 4 | 564 |
| Gnad | 67 | 1979 | 33% to 79% | 5 | 1,459 |
| Jaeger | 21 | 1980 | 30% to 50% | 5 | 787 |
| Shapiro | 104 | 1985 | 21% to 44% | 7 | 900 |
| Catalano | 22 | 1990 | 23% to 44% | 10 | 1,004 |
| Wagner | 107 | 1990 | 21% to 44% | 7 | 972 |
| Hestnes | 72 | 1991 | 23% to 79% | 10 | 2,166 |
| Prasher | 76 | 1994 | 33% to 69% | 6 | 818 |
| Roizen | 23 | 1994 | 27% to 57% | 5 | 266 |
| Berk | 121 | 1996 | 13% to 44% | 7 | 839 |
| Davis | 24 | 1996 | 23% to 44% | 2 | 466 |
| Wong | 135 | 1997 | 12% to 57% | 7 | 507 |
| Haugen | 5 | 2001 | 19% to 70% | 15 | 2,352 |
| Kim | 32 | 2002 | 13% to 57% | 9 | 1,296 |
| Yurdakul | 13 | 2002 | 20% to 38% | 5 | 373 |
| Chan | 136 | 2004 | 20% to 38% | 4 | 481 |
| Ji | 43 | 2006 | 9% to 69% | 10 | 762 |
| Yurdakul | 27 | 2006 | 21% to 44% | 11 | 1,314 |
| Fimiani | 28 | 2007 | 22% to 57% | 7 | 787 |
| Molina | 172 | 2008 | 19% to 42% | 7 | 564 |
| Creavin | 6 | 2009 | 3% to 60% | 20 | 2,286 |
| Kim | 14 | 2009 | 20% to 57% | 6 | 704 |
| Paudel | 140 | 2010 | 12% to 44% | 11 | 1,125 |
| Yahalom | 123 | 2010 | 20% to 50% | 6 | 343 |
| Karlica | 33 | 2011 | 20% to 57% | 5 | 600 |
| Morton | 7 | 2011 | 24% to 46% | 13 | 1,489 |
| Adio | 156 | 2012 | 5.6% to 27% | 7 | 597 |
| Afifi | 8 | 2013 | 6% to 88% | 31 | 3,507 |
| Aslan | 125 | 2013 | 12% to 46% | 6 | 659 |
| Perez | 131 | 2013 | 20% to 60% | 5 | 553 |
| Miyazaki | 15 | 2014 | 20% to 60% | 6 | 585 |
| Ljubic | 88 | 2015 | 20% to 57% | 9 | 1,172 |
| Watt | 16 | 2015 | 19% to 34% | 3 | 269 |
| Nanda | 17 | 2016 | 20% to 57% | 6 | 651 |
| de Weger | 173 | 2019 | 15% to 47% | 5 | 379 |
| Bermudez | 132 | 2020 | 1.9% to 38% | 6 | 1,757 |
| Harrison | 51 | 2020 | 19% to 42% | 5 | 526 |
| Makateb | 9 | 2020 | 5.6% to 65% | 16 | 2,312 |
| Sousa | 36 | 2020 | 30% to 45% | 4 | 713 |
| Mathan | 174 | 2022 | 18% to 57% | 4 | 601 |
| Rojas-Carabali | 134 | 2023 | 9.5% to 38% | 7 | 653 |
| Sun | 19 | 2023 | 9.5% to 57% | 7 | 1,249 |
| current review |  | 2025 | 1.9% to 100% | 137 | 16,781 |

The most frequent ranges are 20-57% (4 times), 21-44% (3 times), 19-42% (2 times), 20-38% (2 times). +, additional reviews were cited, or not-verified sources were quoted

### GLOBAL PREVALENCE and NUMERICAL ESTIMATES

Our analysis established that the global prevalence of strabismus in Down syndrome is 29-39%, depending on ethnicity. When adjusted for ethnic differences, the global prevalence of strabismus in Down syndrome is 30.2% – allowing to estimate the number of Down syndrome people with strabismus at 1.81 million of the presumed 6 million people with Down syndrome.^165^ The large majority of studies examining and reporting the prevalence of strabismus is based on surveys in normal schools.^177^ But many children with developmental disabilities such as Down syndrome or cerebral palsy, to name just two of the most frequent syndromes, do not attend normal schools,^9,40,42,147,154^ and therefore would be excluded in most population-based studies of strabismus. Since children with Down syndrome or with cerebral palsy^178^ have a much larger strabismus prevalence (30-40% – about 15-20-fold higher than in the normal population at 2%),^179^ a substantial number of children with strabismus are missed in the “normal school” based studies, resulting in an undercount.

### AGE AT ONSET

Previous Down syndrome studies reported that the youngest ages (0-3 years) had the lowest strabismus prevalence, while the prevalence increased with age.^4–5,10–20,31,145^ The onset of strabismus (esotropia) in Down syndrome peaks at about 4.5 years of age.^5,15–16,19–20^ The strabismus usually is the acquired type, not congenital, as is most common for esotropia in infants without Down syndrome.^180^ This is consistent with our analysis of studies showing that the strabismus prevalence in Down syndrome at 0-3 years of age is much lower (15.6%) than the prevalence at 15 years and older (53.2%) (Figs. 3C,E, 4A). These data refute the earlier notion that the strabismus in Down syndrome spontaneously resolves in two thirds of cases.^40,65–66,100,181–183^

### CONFOUNDING FACTORS

Besides age and ethnicity, we considered other confounding factors. Since strabismus was diagnosed in nearly all studies by ophthalmologists, differences in methodology are unlikely to be significant. We did not find in the reviewed sources any indication of education or social class having an effect on strabismus prevalence.

### AMBLYOPIA

Opinions about the prevalence of amblyopia in Down syndrome are divided. The majority of studies reported that amblyopia was rare (0-5%)^5,15,23–25,28–31,144^ or “uncommon” (8-14.3%),^7,21–22,26,96,100,114,160^ while a smaller number of authors found a relatively large prevalence of amblyopia (16.9-36.4%)^10,12–13,27,38,109,174,184^ and one study reported 54.5%.^134^ However, only a relatively small fraction of the amblyopia cases associated with strabismus, meaning that the majority of amblyopia cases in Down syndrome was not caused by strabismus,^17,25–27,109,113,127,160^ but some appear to be caused by anisometropia^25,113^ which is more frequent in older children with Down syndrome.^11,21,80,174^ The overwhelming consensus is that, probably due to the late onset of strabismus, binocular vision is often preserved in Down syndrome.^5,15,23–31^

### GENDER

There were slightly more males than females in our cohorts (sex ratio of 1.21:1), consistent with previous reports.^55–56,166^ The bias towards males presumably is due to genetic mechanisms (joint segregation of chromosomes 21 and Y).^166^ Similar to a previous analysis of gender in strabismus within the normal population,^185^ we found no gender difference in the prevalence of strabismus in Down syndrome. There are slightly more males than females with strabismus in Down syndrome, but only because there are slightly more males than females with Down syndrome.

### ET/XT RATIO

Ethnic differences in the esotropia/exotropia ratio were noted by several investigators.^14,17,33–39^ Caucasians with Down syndrome have a further increased bias towards esotropia, with exotropia being rare, while ethnicities that normally have more exotropia than esotropia also have a bias towards esotropia when Down syndrome is present, but the esotropia/exotropia ratio is much lower (3.21) than in Caucasians with Down syndrome (9.98). The likely explanation for these ethnic differences is the difference in the orbital anatomy where the orbital width is already narrow in Caucasians, but becomes even more narrow in Down syndrome as discussed in more detail below (“Orbital Anatomy”).

### ETIOLOGY OF STRABISMUS

Authors noted that strabismus in Down syndrome differs substantially from that in children without the syndrome,^19,49–51^ but the reason(s) have remained enigmatic. While some authors state that the etiology of strabismus in Down syndrome is unknown,^19,21,51,151^ several possibilities have been discussed.^5,14,19,21,37,40–42,44–45,47–49,65,173,176,186^ We will review them in a historical (chronologic) sequence and discuss their merits.

Orbital Anatomy. The inclination of the orbit is abnormal and the width of the orbit is much more narrow in Down syndrome.^42,99,176,187–189^ Indeed, the interpupillary distance as a measure of the orbital width is reduced in Down syndrome, by about 5-10 mm in Caucasians when compared to age-matched children or adults without Down syndrome,^3–4,21,40–42,48,64,99,146,187,190–193^ and by 1-3 mm in Asians and West Africans.^156,193^ For geometric-mechanistic reasons, first described in the late 19^th^ century, optimal extraocular muscle function and binocular vision require an orbital anatomy that is within certain normal limits. When the eyes are too narrow set, or too wide set, as in Down syndrome for the narrow extreme and craniosynostosis for the wide extreme,^5,21,41,124,193–195^ then esotropia or exotropia are much more likely, because the medial or lateral rectus muscles operate in a suboptimal frame.^21,41,196–205^ The ethnic differences in the ET/XT ratio in Down syndrome are consistent with the notion that orbital anatomy and especially short orbital width and a narrow interpupillary distance are a major contributing factor in the etiology of strabismus in Down syndrome.

Muscle hypotonia. Hypotonia of skeletal muscles in Down syndrome is well known.^22,44,51,55–56,95–96,170^ Whether this applies to the extraocular muscles is unclear.^175^ The “excessive power of the medial rectus muscles” was noted,^40^ which contradicts the idea of hypotonia causing strabismus in Down syndrome.

Lens opacities/ cataract. The prevalence of cataracts is increased in Down _syndrome, especially in older individuals._^6,10,13,18–19,21,24,29, 38,43,55,57,69,71–72,82,103,105,127,142,148^ Some authors proposed that lens opacities are associated with, and may be a cause of, strabismus in Down syndrome.^40,42,65,71^ However, cataracts typically develop in older children, while congenital cataracts are relatively rare in Down syndrome.^4,7–8,10,17,23,27,30–33,40,64,66,68,83–84,88,90,92,99,109,114,131,135,140,145,147,150,172,186^ Thus, there is a mismatch between the onset of significant lens opacities and the peak onset of strabismus in Down syndrome, indicating a minor, if any role for cataracts in the etiology of strabismus in Down syndrome.

Refractive errors. Refractive errors are the most common ocular defects in Down syndrome. Early studies implicated them as a potential cause of strabismus.^37,40,42–43,47,65–66^ Most studies report more hyperopia than myopia in Down syndrome,^4–6,8,11,13–14,16–17,27–28,31–32,34–35,39,42–43,57,59–61,64,67,69,77–78,84,86,88,90–91,114,121,127,129,133–135,138,140,143,146,148–149,151,153–154,157,160,172,184,186^ but about one quarter of the studies (26.4%) report more myopia than hyperopia.^15,18,21,23,26,33,63,65,72,81–82,94,105,113,119,136,141,150^ Some authors stated that in Down syndrome, hyperopia is more common in Caucasians, and myopia is more common in Asians.^8,19,37,39^ However, in the strabismus cohorts we compiled, hyperopia was about three times more frequent than myopia, in both Caucasians and Asians – more hyperopia was reported in most Asian studies,^14,17–18,32,35,135,138,140,143,146,148-149,151,153^ while more myopia was reported in fewer Asian studies.^18,136–137,141,150^ In normal children with refractive errors, the hyperopia can resolve over time (emmetropization), but such emmetropization does not occur in children with Down syndrome. This has been called a failure of emmetropization and has been implicated as a possible cause of strabismus in Down syndrome.^11,16,19,37,43,45,47,49,140,206^ However, in contrast to normal children,^206^ there does not seem to be any convincing association between either hyperopia or myopia and development of strabismus in Down syndrome.^11,14,45,49,51,86,105,208^

Astigmatism is one of the most frequent ocular findings in Down syndrome.^7–8,10–11,14,16,35–37,43,45,63,87,104,127,144,174,208–209^ Similar to the failure of emmetropization, astigmatism does not decrease with age in Down syndrome.5,11,38-39,174 However, astigmatism does not appear to be associated with strabismus in Down syndrome.5,11,49,87,134

Accommodation weakness. Studies noted that the esotropia in Down syndrome often has an accommodative component.^21–22,24–25,40,42,67,96^ Children with Down syndrome fail to develop an adequate accommodative convergence mechanism – an abnormality of accommodation that was subsequently described in more detail.^5,7,11,15-16,19,37–39,43,48,115,138,141,151,174,206,208–215^ The possibility of this deficit being due to mechanics of a thinner cornea and lens and reduced lens power has been discussed.^16,174,209–210^ Other possibilities include sensory pathway deficits,^216^ peripheral motor abnormalities (ciliary muscle), or central abnormalities (neuronal control of the ciliary muscle).^59,208–210,213,217^ The ciliary muscle appears normal in Down syndrome.^16^ Potential defective neural control of accommodation may be a manifestation of a general cholinergic deficit in Down syndrome.^218^ The interplay of accommodation weakness, hyperopia, and a decreased interpupillary distance in Down syndrome may precipitate strabismus.^219^ Use of bifocals improves accommodation accuracy, near visual acuity, and reduces the degree of deviation in small-angle esotropic Down syndrome children.^173,214,221,222^

Visual acuity and contrast sensitivity. Several studies revealed deficits in visual acuity and contrast sensitivity in Down syndrome that developed after one year of age.^111,216,223–225^ These deficits may be due to pre-retinal (optic) abnormalities, they could be cortical, a consequence of accommodation weakness, or a combination of the above factors.^16,37,111,216,223,226–227^ The cornea and lens are thinner in Down syndrome, as mentioned.^16,174,208–210^ Although the fovea of the retina has an abnormal thickness and layering in Down syndrome and also in animal models of Down syndrome,^46,228–229^ a thicker macula does not seem to correlate with reduced visual acuity.^228^ Regarding a possible cortical contribution, the visual cortex of Down syndrome children has age-related abnormalities in the neuronal architecture, with reduced dendritic arborizations and reduced neuronal densities,^16,37,230–236^ as well as slightly reduced synaptic density (by 1-9% at ages 4-9 years),^234^ but such reports await confirmation with modern stereological methods.^237^ Recent analyses concluded that there are no profound disruptions in synaptic formation and/or pruning in Down syndrome.^238^ While a lesion of inputs to visual cortex can cause strabismus,^239^ it is unclear to what extent subtle cortical abnormalities in Down syndrome (lacking any acute injury) may contribute to reduced binocular vision processing and the development of strabismus. On the basis of exotropia being more frequent than esotropia in cases of brain damage, Haugen and Hovding^5^ argued against a major contribution of cortical abnormalities causing strabismus in Down syndrome. Also, there is no direct evidence showing that the minor visual cortex abnormalities in Down syndrome contribute to strabismus.

Combination of sub-normal conditions. Multiple factors rather than a single factor along the sensory-motor loops (optic apparatus, retina, visual cortex, oculomotor nuclei, extraocular muscles, orbital anatomy)^240–241^ may be necessary to elicit strabismus in Down syndrome. Given the ethnic differences in the esotropia/exotropia ratio and their correlation with orbital anatomy, the orbital width appears to be a major factor.

## Conclusion

A large number of people with Down syndrome have strabismus: about 1.81 million, with a lifetime occurrence of 3.19 million. There are major ethnic differences in the esotropia/exotropia (ET/XT) ratio, with Caucasians having a high ET/XT ratio (9.98), while this ratio is much lower in other ethnicities (3.21). Surprisingly, the abnormal orbital anatomy in Down syndrome is rarely considered as contributing or being a major cause of strabismus, even though an abnormal width of the orbit is a known risk factor for strabismus.^41,194–195,198,204^ Ethnic differences in the ET/XT ratio support the notion that orbit differences substantially contribute to the etiology of strabismus in Down syndrome. A combination of retinal differences, cortical abnormalities, accommodation weakness, blurry vision, together with abnormally narrow orbital width produces multiple conditions of sub-normality in Down syndrome that may prevent the development of normal binocular processing.^42,242^

## Data Availability

All data produced in the present work are contained in the manuscript and the supplementary material

## Acknowledgments

We thank Jenny Costa (University of Nevada, Reno, School of Medicine) for her help with retrieving sources. We also thank Michael Herron and Mitchell Strominger (University of Nevada, Reno School of Medicine), Zainab Zehra (COMSATS University, Islamabad, Pakistan), and Wei Yang (University of Nevada, Reno School of Public Health) for helpful comments.

## SUPPLEMENTARY MATERIAL

**Supplementary Table 1.**
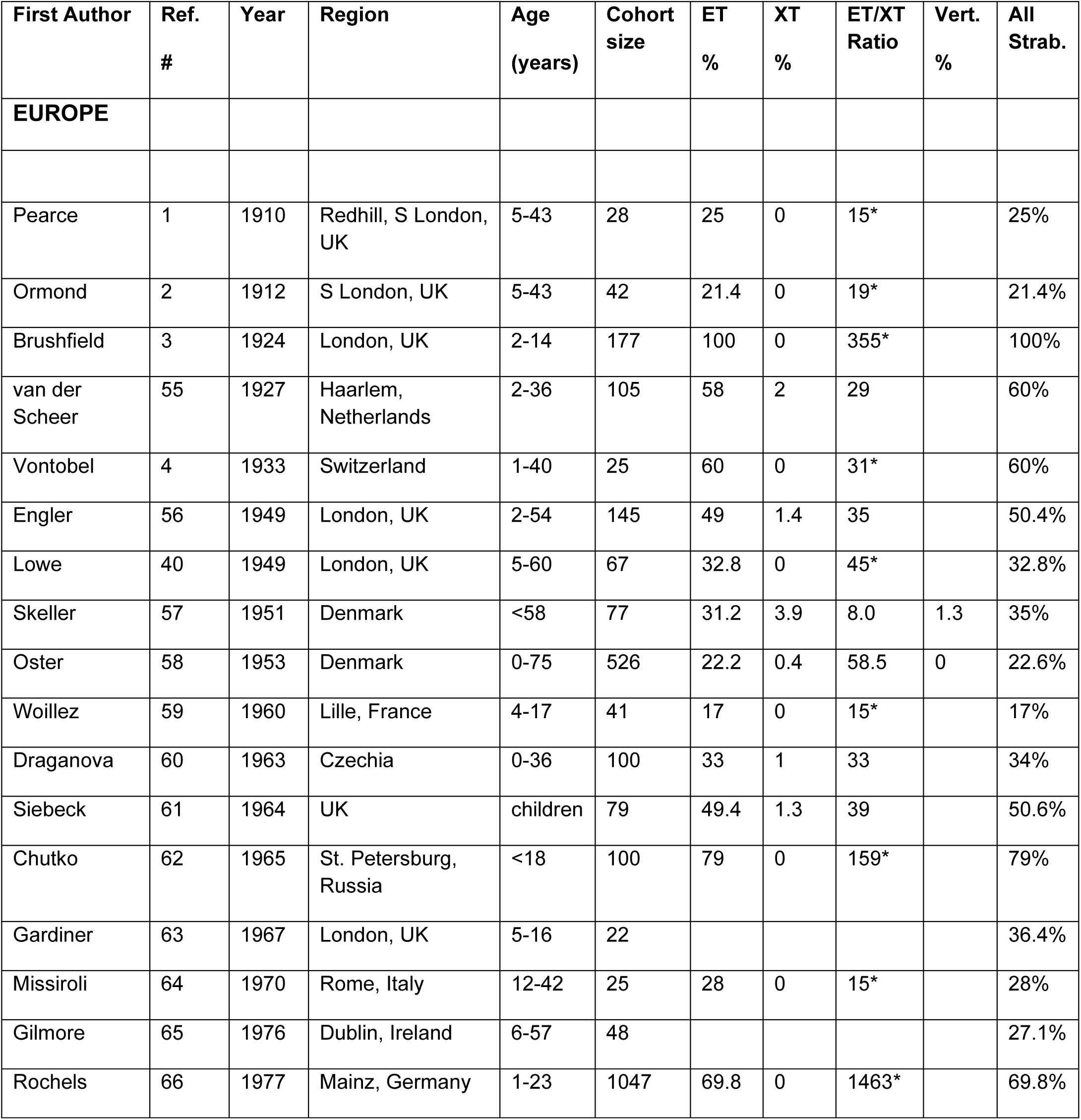
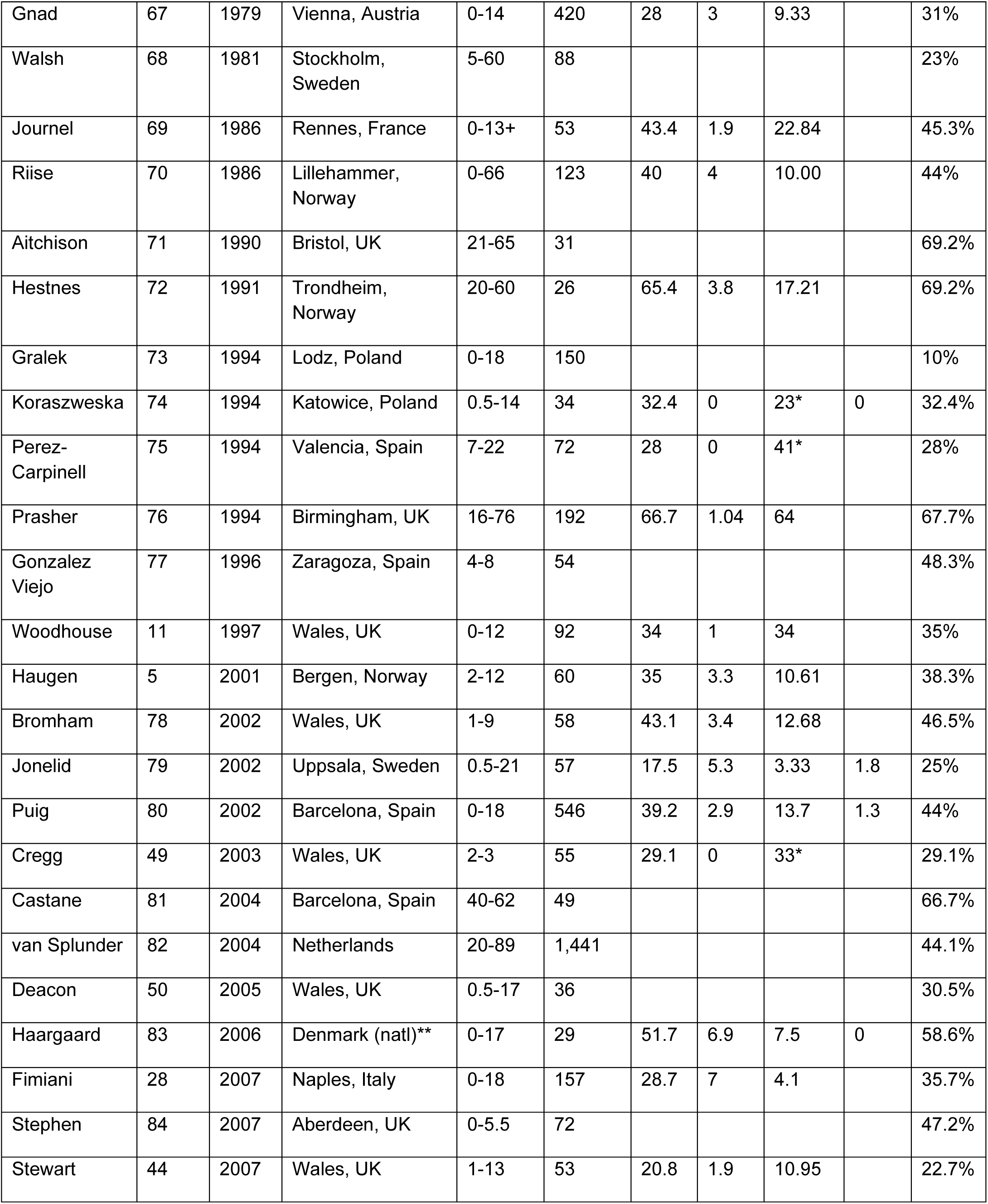
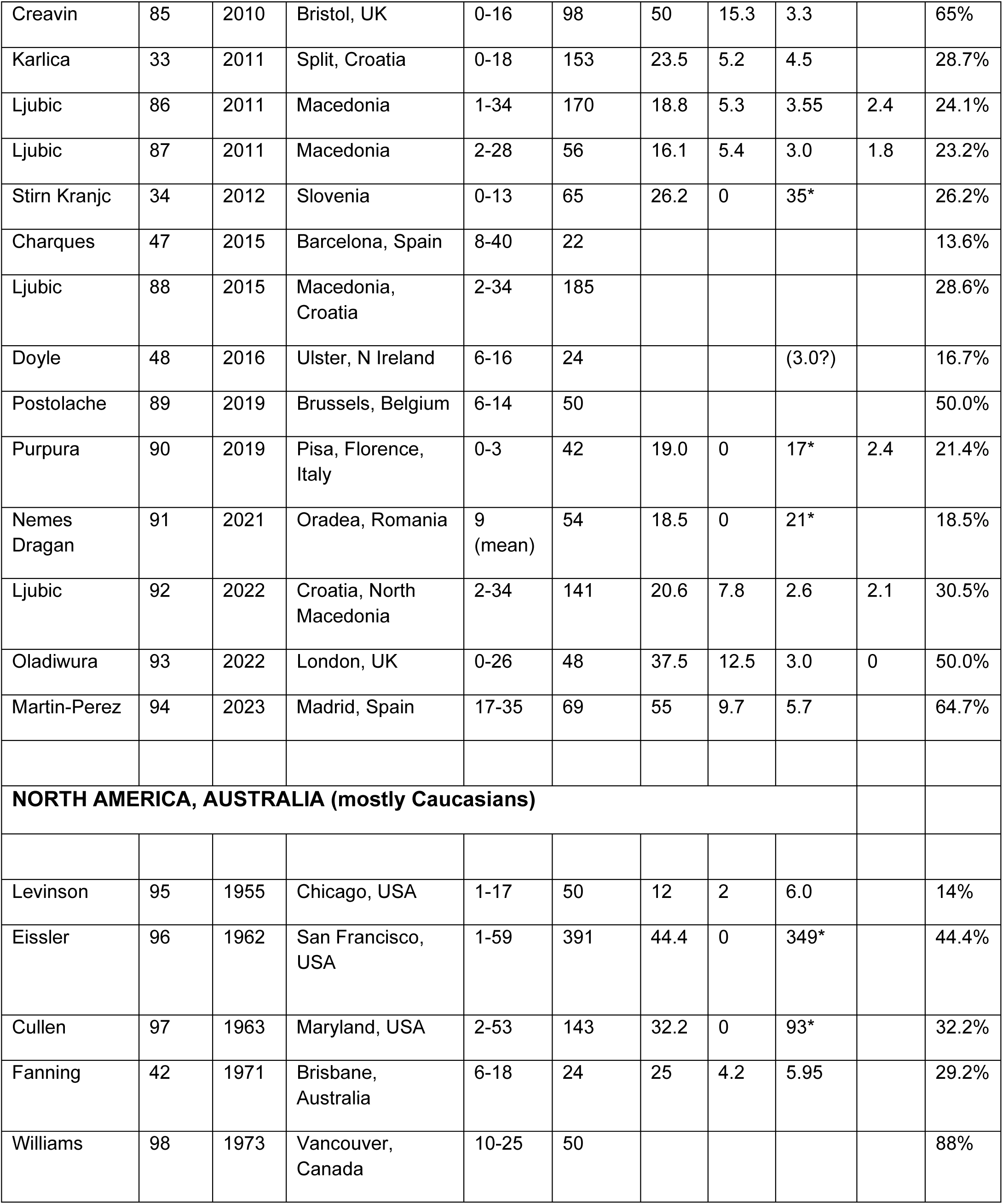
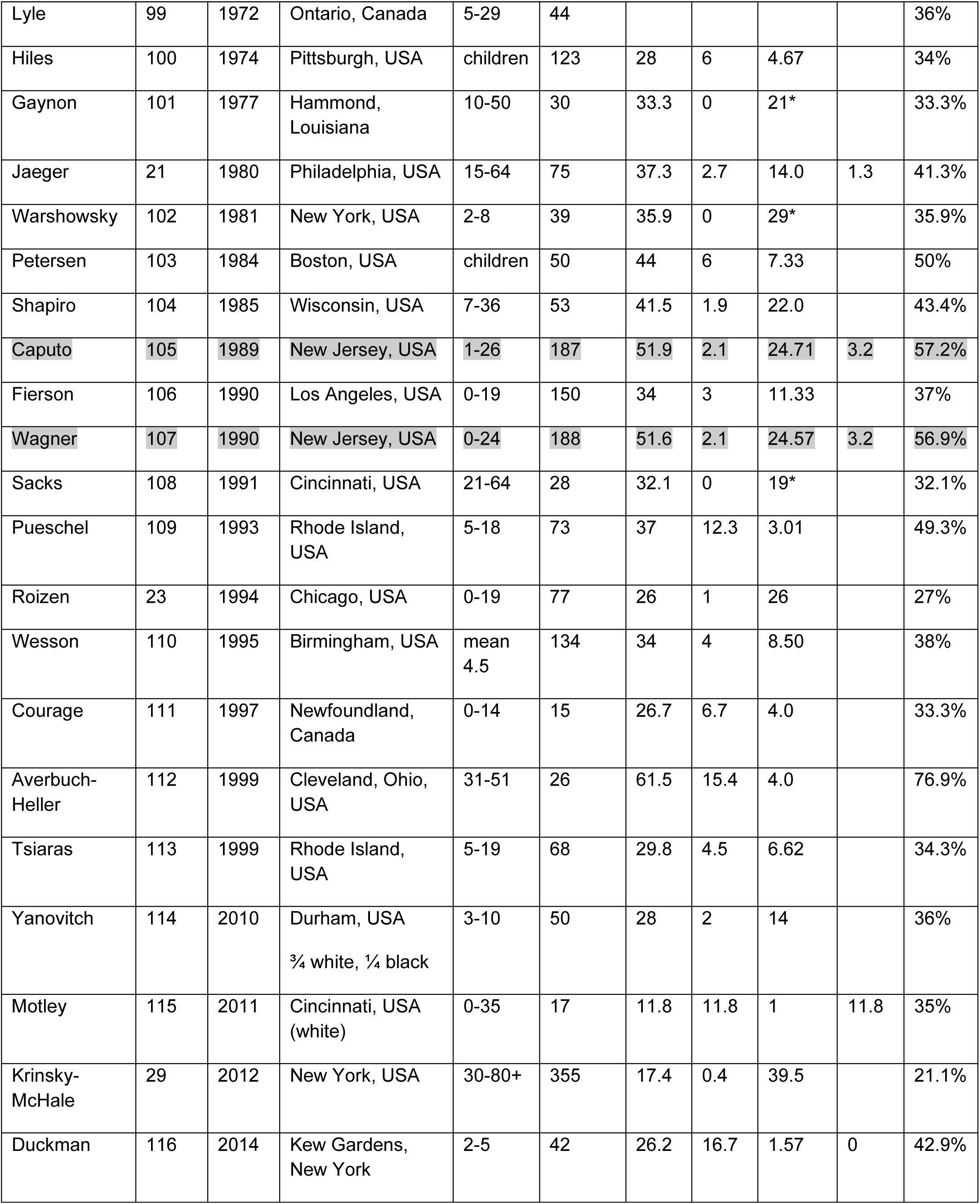
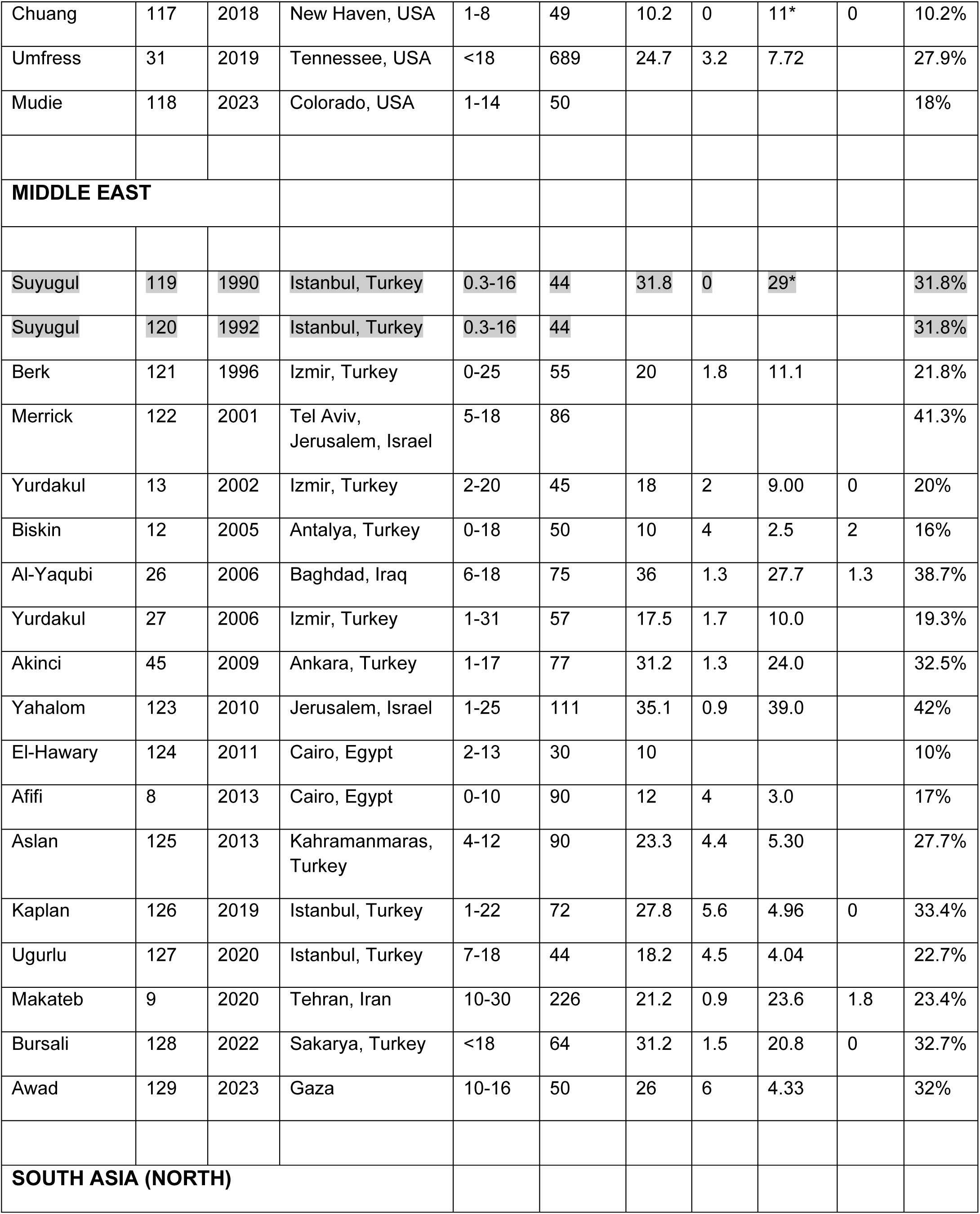
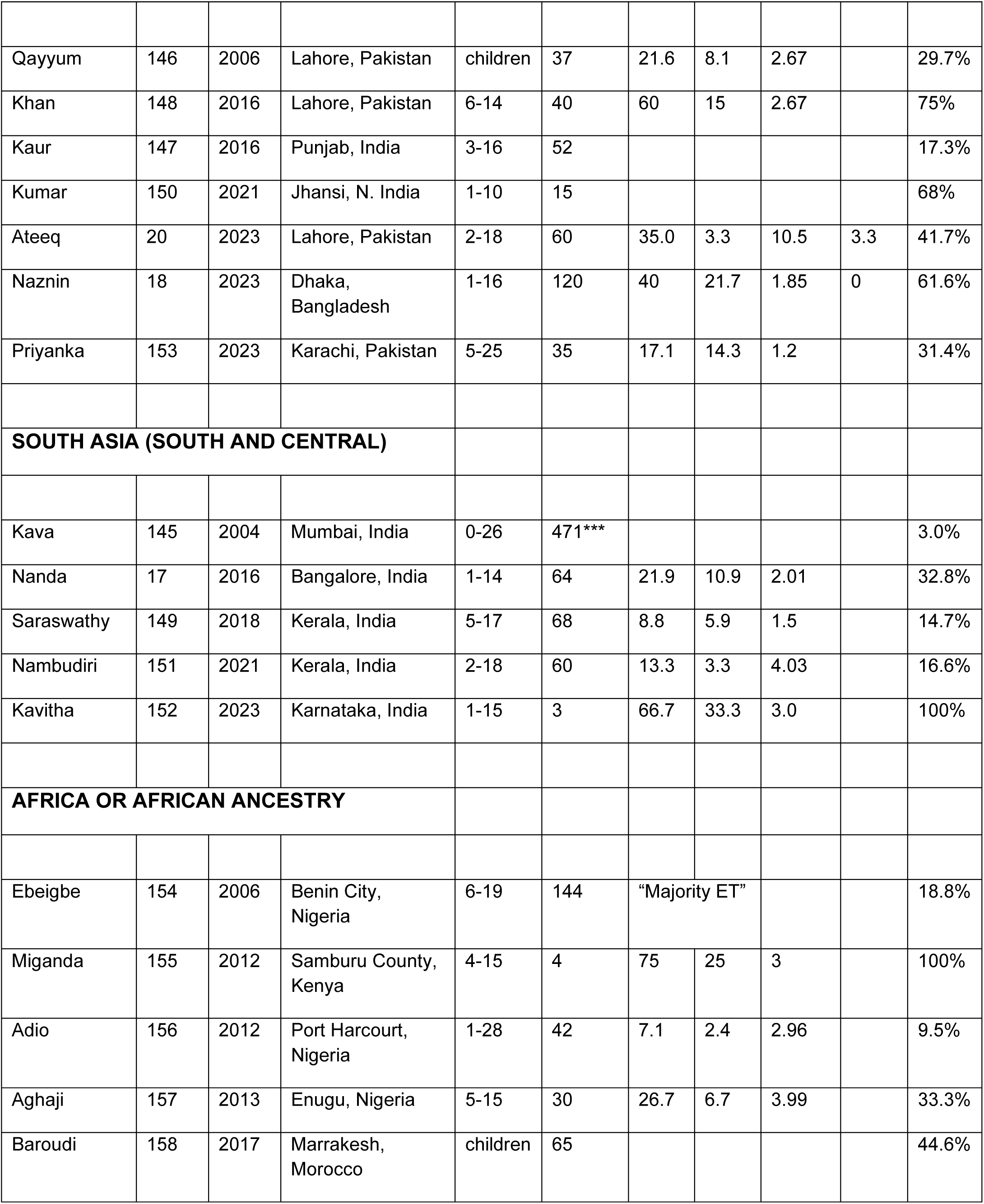
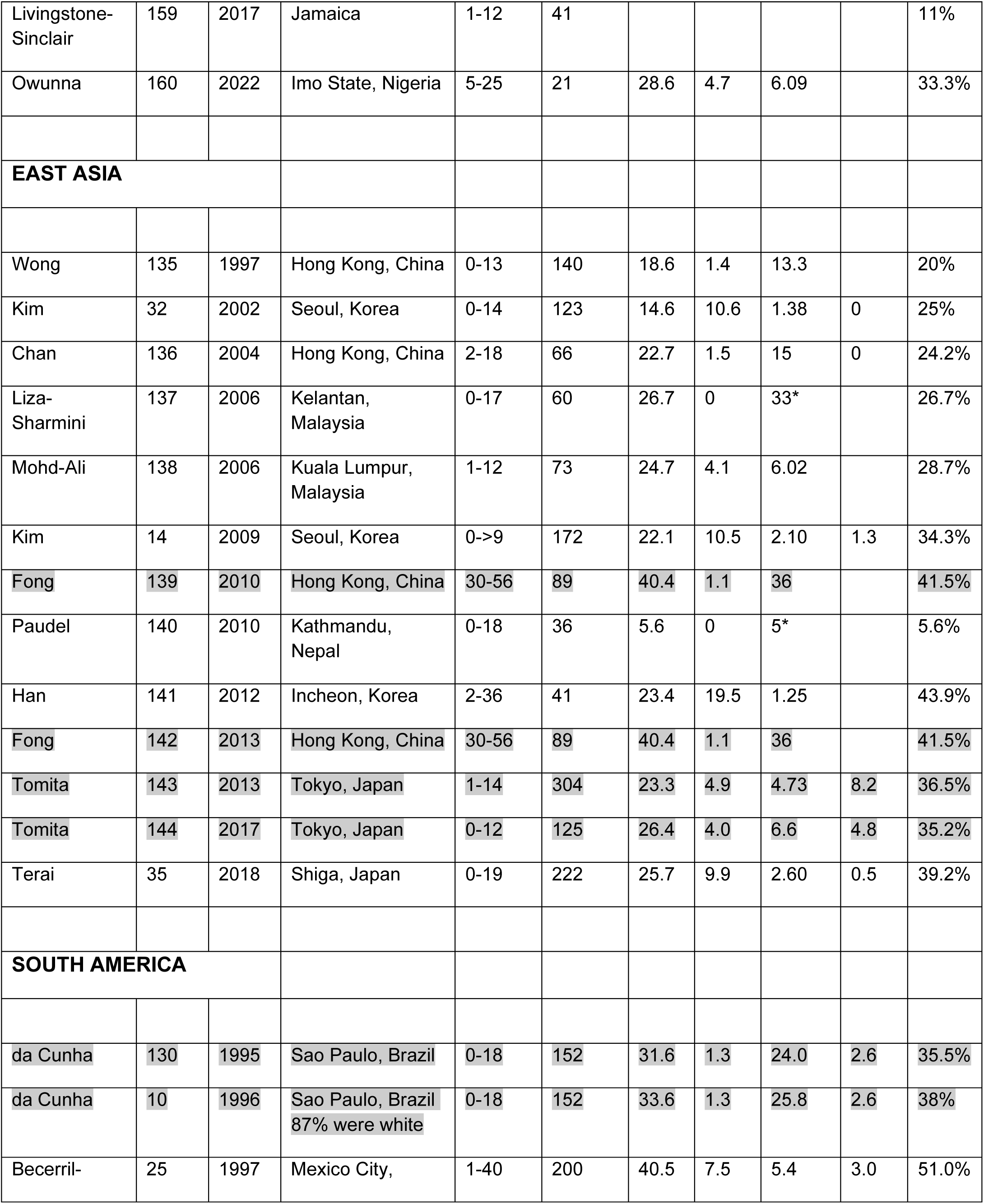
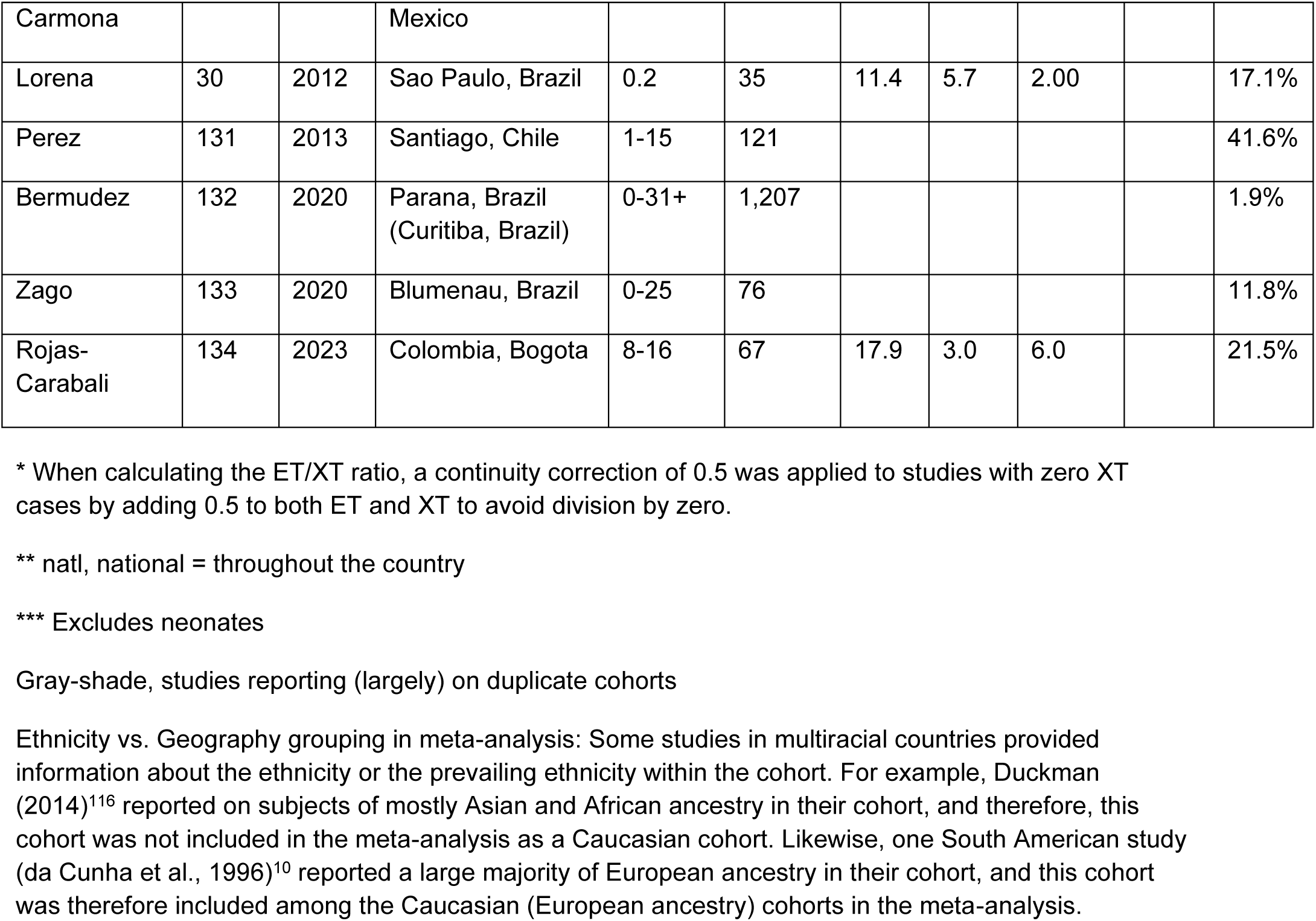
Prevalence of strabismus, esotropia (ET), exotropia (XT), vertical deviation (Vert.) and all strabismus (All Strab.) in cohort studies of Down syndrome. Ref., Reference. The Table is organized by region/ethnicity as well as chronological within those regions. Explanations: +, a few subjects in the cohort exceeded the range; blank, no information was given. N=142 studies; shaded: duplicate reports of the same cohort.

**Supplemental Fig. 1.**
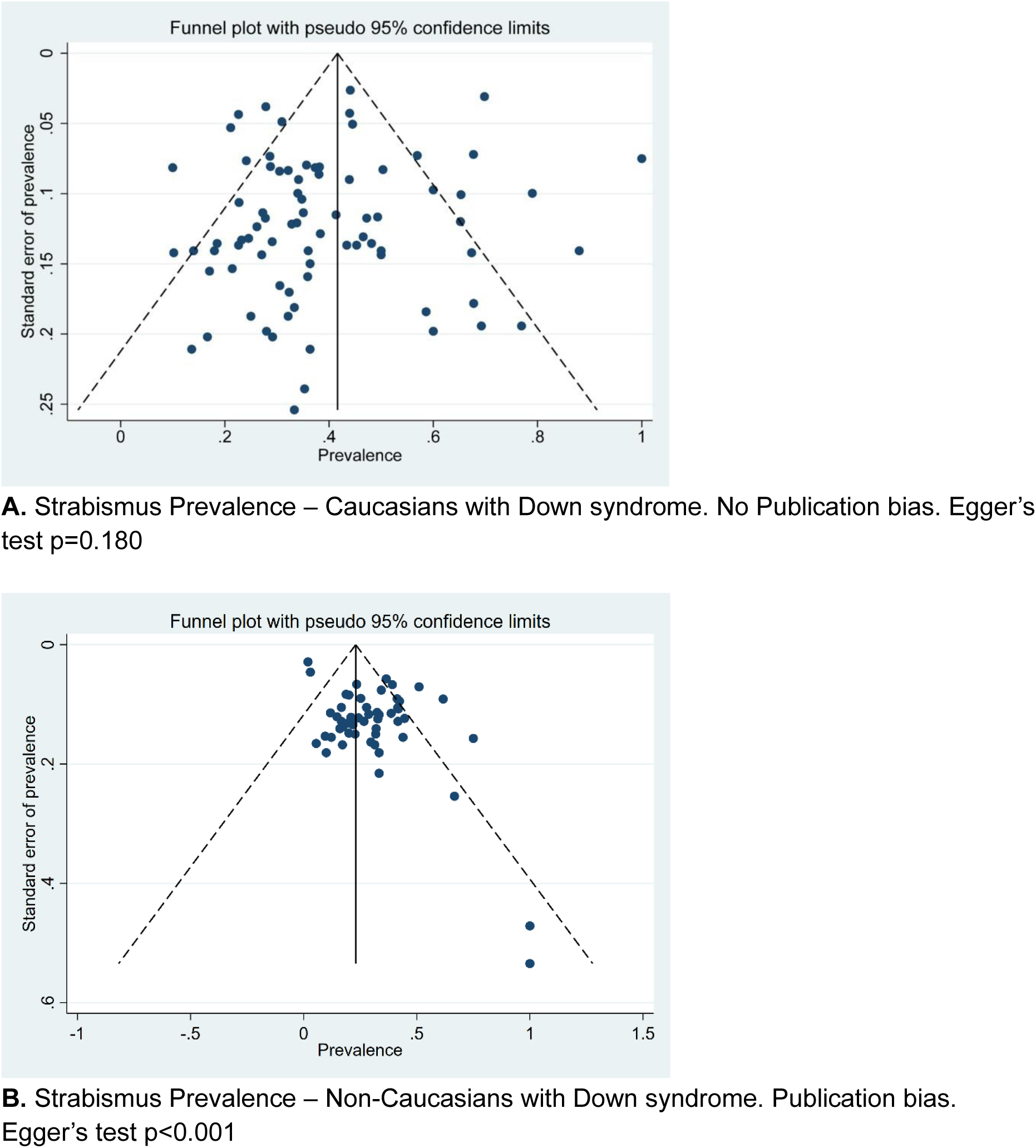
**A,B.** Funnel Plots for the prevalence of strabismus in Down syndrome in Caucasians (**A**, n=84 studies) and for Non-Caucasians (**B**, n=53 studies).

**Supplemental Fig. 2.**
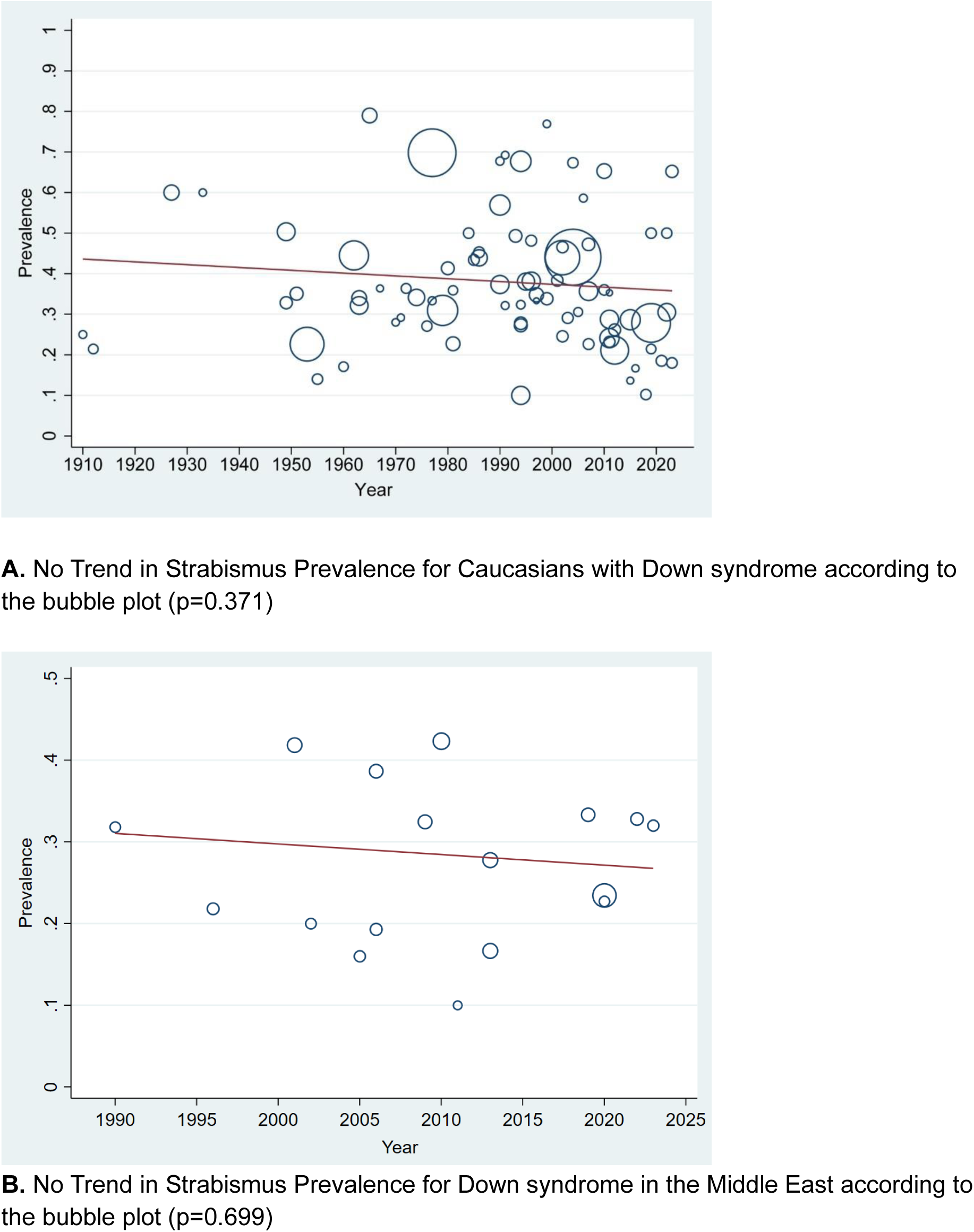
**A,B.** Bubble plots showing Longitudinal Analysis for Trends in Strabismus Prevalence for people with Down syndrome in Caucasians (**A**) and in the Middle East (**B**): no generational changes. We identified two outliers in studies on Caucasians (Brushfield, 1924; Williams et al., 1973)^3,98^ using Tukey’s Hinges with k=1.5. These outliers were excluded from the longitudinal trend analysis.

**Supplemental Fig. 3.**
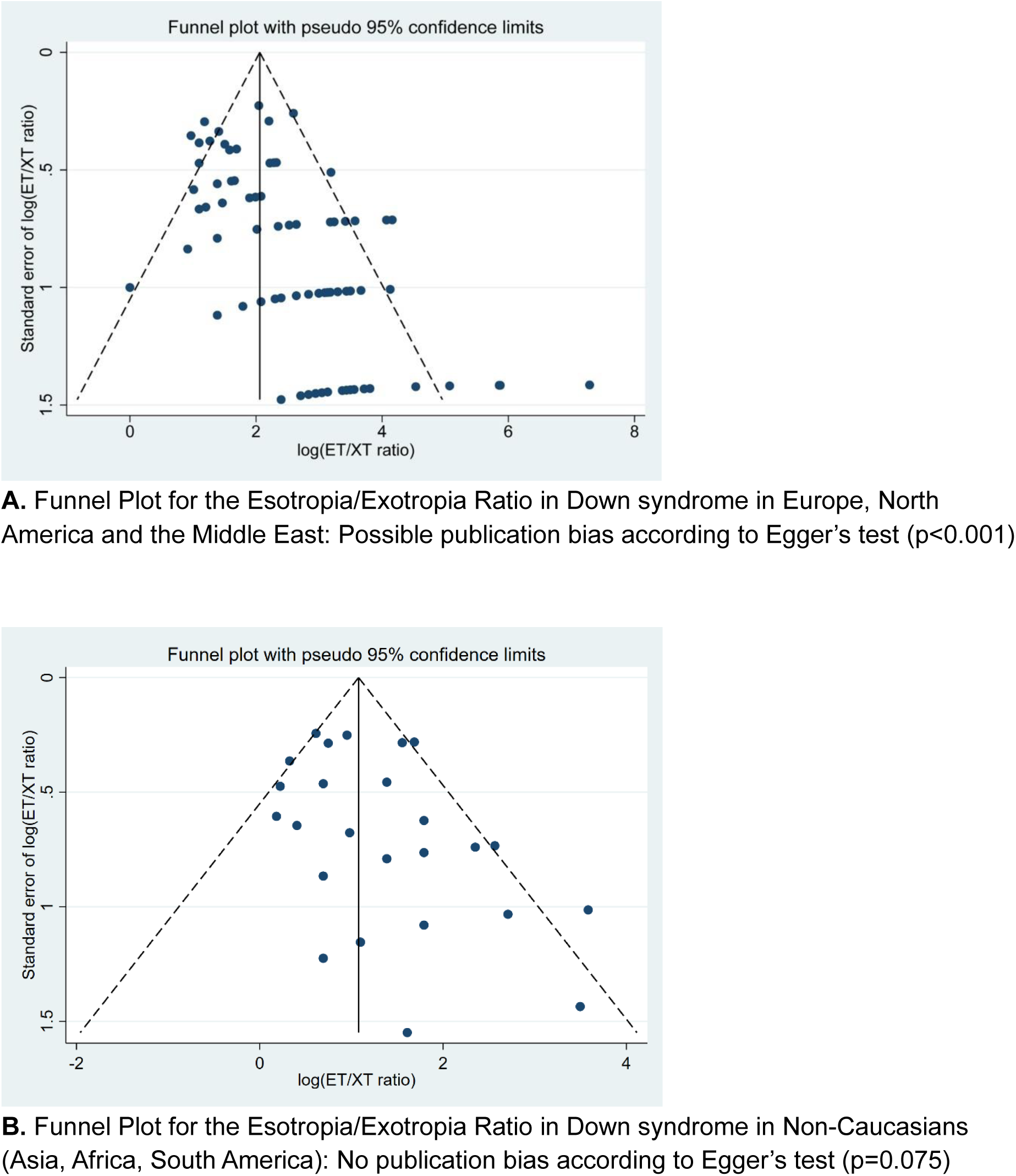
**A,B.** Funnel Plots showing possible publication bias in the Esotropia/ Exotropia Ratio in Caucasians with Down syndrome (**A**), but not in Non-Caucasians (**B**).

**Supplemental Fig. 4.**
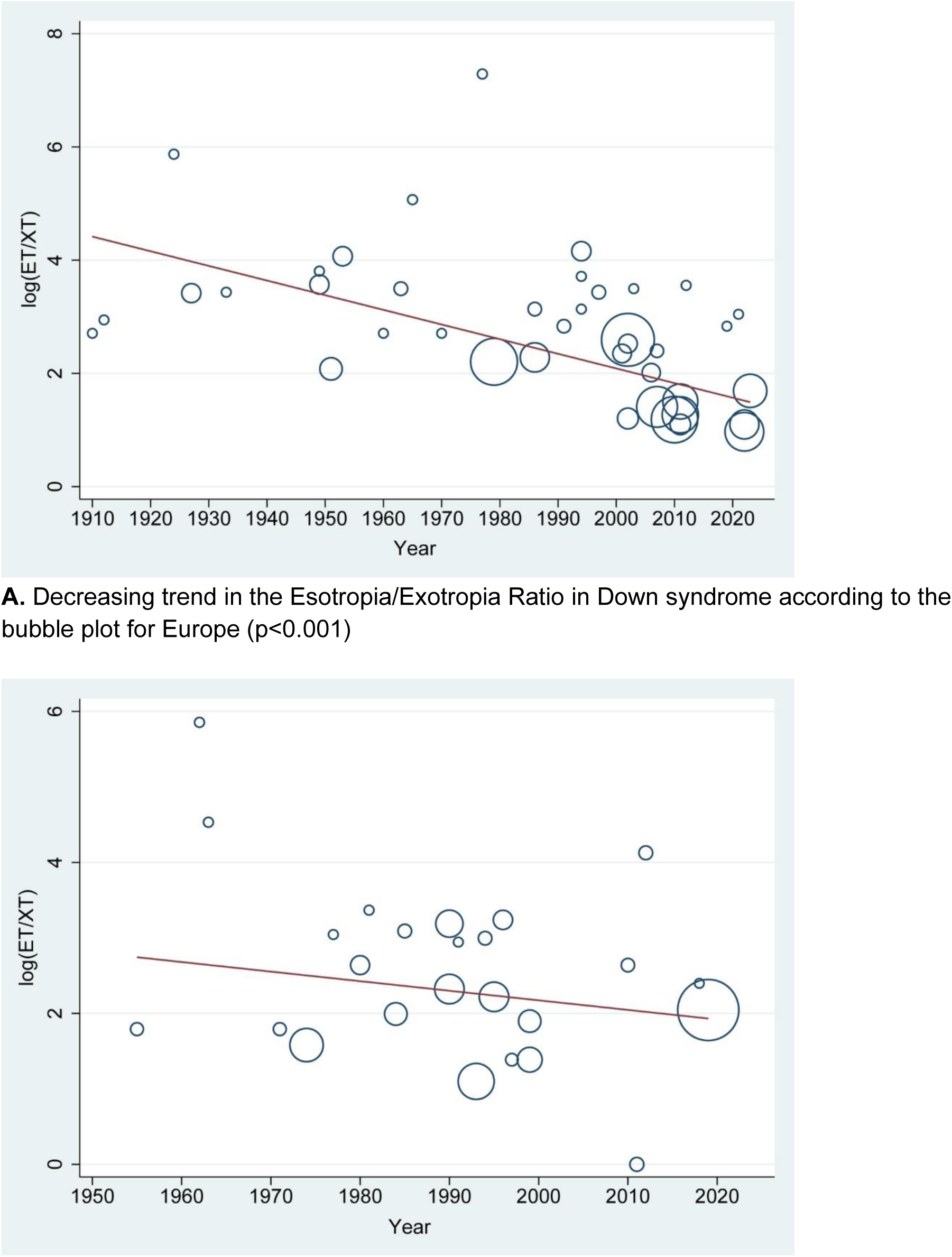
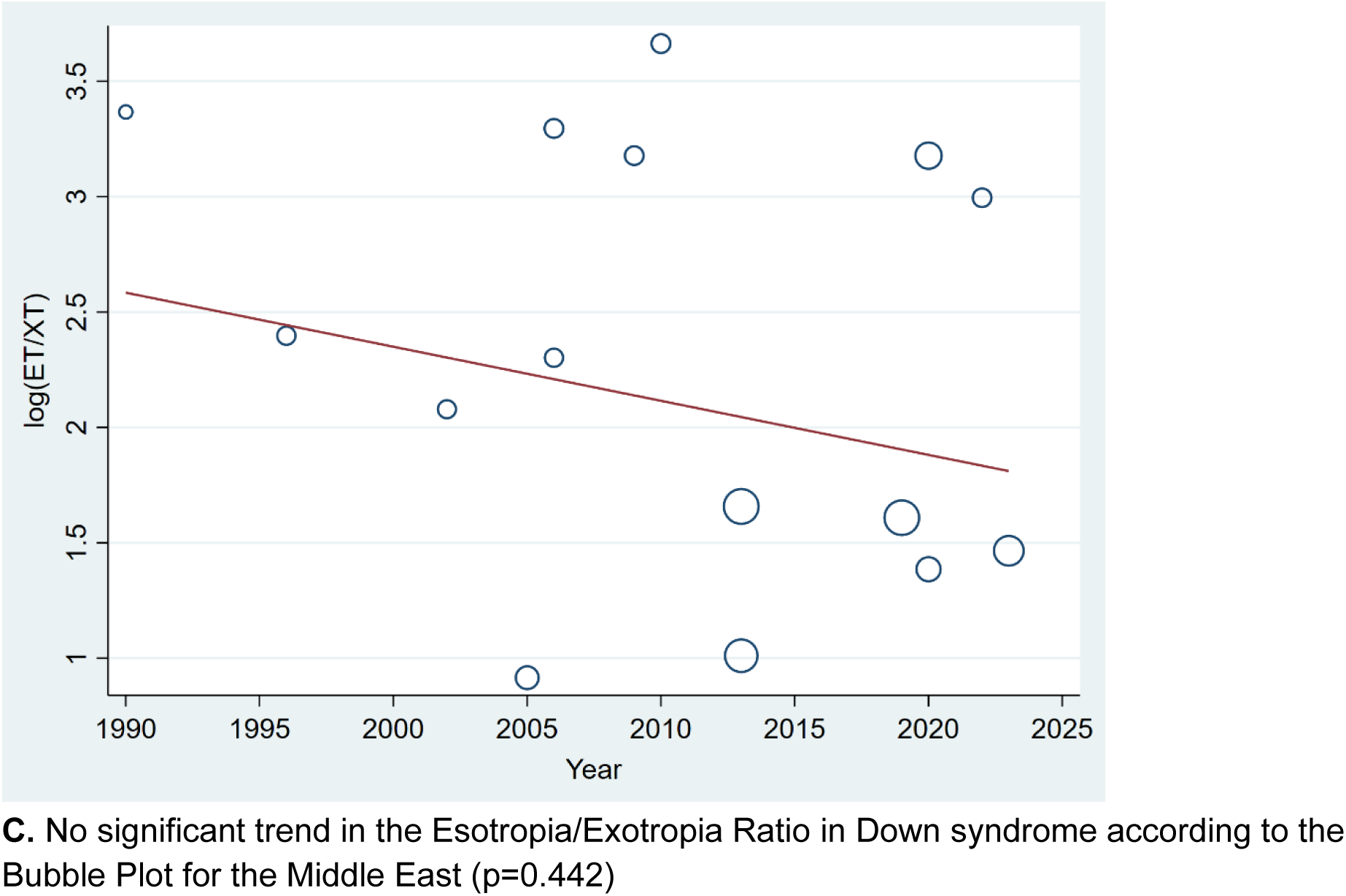
**A-C.** Bubble Plots showing Longitudinal Analysis for Trends in the Esotropia/Exotropia Ratio in Down syndrome: Generational changes in Europe (**A**), but not in North America (**B**) or in the Middle East (**C**).

